# Clinical consequence of vessel perforations during endovascular treatment of acute ischemic stroke

**DOI:** 10.1101/2023.06.02.23290912

**Authors:** P. Matthijs van der Sluijs, R. Su, S.A.P. Cornelissen, A.C.G.M. van Es, G. Lycklama a Nijeholt, B. Roozenbeek, P.J. van Doormaal, J. Hofmeijer, A. van der Lugt, T. van Walsum, MR CLEAN Registry investigators

**Author notes:** **Corresponding author:** P. Matthijs van der Sluijs, Department of Radiology and Nuclear Medicine, Erasmus MC University Medical Center, Rotterdam, The Netherlands, Room Na-2513, 3015 CE, Rotterdam, The Netherlands. Full list of contributors is listed in the supplemental appendix.

## Abstract

**Background:** Endovascular treatment (EVT) of acute ischemic stroke can be complicated by vessel perforation. We studied the incidence and determinants of vessel perforations. In addition, we studied the association of vessel perforations with functional outcome, and the association between location of perforation on DSA and functional outcome, using a large EVT registry.

**Methods:** We included all patients in the MR CLEAN Registry who underwent EVT. We used digital subtraction angiography (DSA) to determine whether EVT was complicated by a vessel perforation. We analyzed the association with baseline clinical and interventional parameters using logistic regression models. Functional outcome was measured using the modified Rankin Scale (mRS) at 90 days. The association between vessel perforation and angiographic imaging features and functional outcome was studied using ordinal logistic regression models adjusted for prognostic parameters. These associations were expressed as adjusted common odds ratios (acOR).

**Results:** Vessel perforation occurred in 74 (2.6%) of 2794 patients who underwent EVT. Female sex (aOR 1.9 [95%CI 1.2-3.1]) and distal occlusion locations (aOR 2.2 [95%CI 1.4-3.7]) were associated with increased risk of vessel perforation. Functional outcome was worse in patients with vessel perforation (acOR 0.41, 95%CI 0.25-0.68) compared to patients without a vessel perforation. No significant association was found between location of perforation or successful reperfusion and functional outcome.

**Conclusion:** The incidence of vessel perforation during EVT is low, but it has severe clinical consequences. Females and patients with a distal occlusion locations are at higher risk. The location of vessel perforation or successful reperfusion did not affect functional outcome.

## Introduction

Endovascular treatment (EVT) in acute ischemic stroke has been demonstrated to improve functional outcome.^1^ Complications such as clot embolization, vessel dissection or vessel perforation are rare, but can have significant negative impact on clinical outcome.^2-4^ in particular, vessel perforation has substantial negative clinical impact.^2^ A perforation of the vessel during EVT causes extravasation of blood and visible contrast material outside the contours of the normal vessel on DSA. Identifying determinants of vessel perforation might help reducing the occurrence. Secondly, identifying angiographic features of vessel perforation, such as anatomical location or current reperfusion status, that are associated with clinical outcome could influence the decision whether to halt or continue EVT.

In several randomized clinical trials, the incidence of vessel perforations was 0.8-4.9%.^5–9^ Two cohort studies describing poor clinical outcomes of vessel perforations, reported an incidence in respectively 16/1599 (1%) and 32/1419 (2.3%) patients.^4,10^ The limited amount of cases in these studies hampers robust assessment of determinants of vessel perforation. In our study, we aimed to address this using a large EVT registry.

## Methods

### Study design and patient selection

We used data from the MR CLEAN Registry, a prospective observational study of all anterior and posterior circulation acute ischemic stroke patients who underwent EVT in the Netherlands.^11^ We used data collected from March 2014 until November 2017. We excluded patients in whom intracranial access was not achieved or in whom a diagnostic DSA was performed solely because of early reperfusion. (Supplemental Figure 1). This study is reported in accordance with the STROBE guidelines (Strengthening the Reporting of Observational Studies in Epidemiology).^12^

### Angiographic analysis and definitions

All imaging in the MR CLEAN Registry was scored by an experienced core laboratory, while being blinded to all clinical data except symptom side. We analyzed the DSAs of vessel perforations reported by interventionists or imaging core laboratory. In addition, we analyzed DSA images of patients who developed intracranial hemorrhage on follow-up CT. In the MR CLEAN Registry, patients received follow-up CT when deteriorated clinically after EVT.

Two experienced neuro-interventionists (observers) analyzed the DSAs separately, and disagreement between observers was resolved by consensus. We annotated the anatomical location, type, and self-limiting contrast extravasation. Anatomical locations were divided into proximal and distal locations. Proximal locations consisted of intracranial carotid artery (ICA), intracranial carotid artery top (ICA-T), M1, A1, and P1 segments. Distal locations consisted of M2, M3, M4, A2 and P2 segments. To distinguish between M1 and M2 segments, we defined the post bifurcation branches of M1 as M2 segments. The contrast extravasation was subdivided into three types: type 1 represents subarachnoid contrast extravasation into the subarachnoid space, type 2 represents contrast extravasation in the adjacent parenchyma of the vessel, and type 3 represents contrast extravasation into the adjacent vein or dural sinus, which leads to an arterio-venous fistula (Figure 1). Successful reperfusion was defined as eTICI 2B-3.^13^

**Figure 1.**
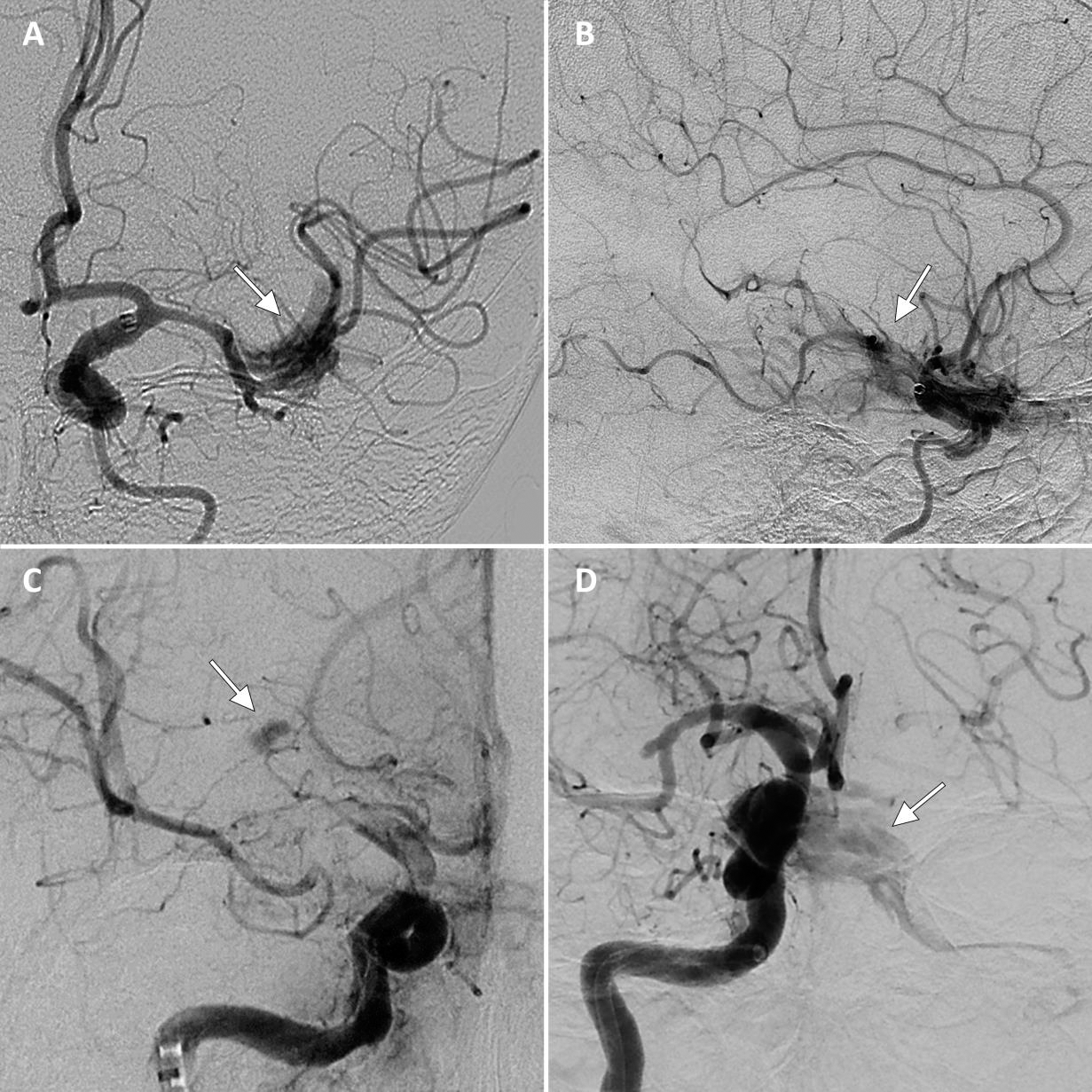
Vessel perforation types based on contrast extravasation on digital subtraction angiography. Arrows indicate contrast extravasation. **A.** Anteroposterior view of subarachnoid type contrast extravasation in the M2 segment **B.** Lateral view of subarachnoid type contrast extravasation in the M2 segment **C.** Anteroposterior view of parenchymal type contrast extravasation surrounding lenticulostriate arteries of the M1. **D.** Lateral view of AV-fistula type contrast extravasation due to occurrence of carotid cavernous fistula. AV fistula, arteriovenous fistula; ICA, internal carotid artery; M(segment), middle cerebral artery;

### Outcome measures

The primary outcome measure was functional recovery according to the modified Rankin Scale (mRS) determined at 90 days.^14^ This ordinal scale ranges from 0 to 6, in which 0 represents ’no symptoms’ and 6 represents ’death’. Good functional outcome was defined as mRS 0-2. The secondary outcomes were the National Institutes of Health Stroke Scale (NIHSS) score at 24-hour follow-up, and 90-day mortality.

### Statistical analysis

Binary logistic regression was used to assess determinants of vessel perforations. The independent determinants were the baseline demographics. Determinants with p < 0.1 in the univariable regression were included in the adjusted regression models.

Ordinal logistic regression, binary logistic regression and linear regression was used to estimate the association of a vessel perforation with mRS at 90 days, with good (mRS 0-2) functional outcome or mortality, and with NIHSS at 24-hour follow-up, respectively. The NIHSS at 24-hour follow-up was log transformed to meet the assumptions of normally distributed residuals for linear regression. The regression models for clinical outcome were adjusted for prognostic patient and interventional characteristics: age, NIHSS baseline, pre-stroke mRS, diabetes mellitus, previous stroke, systolic blood pressure, glucose, intravenous thrombolytics, ASPECTS, occlusion location dichotomized (proximal, distal), collaterals, time from stroke onset to groin puncture, and dichotomized reperfusion status (eTICI≥2B).^15^ In four cases we could not identify the contrast extravasation on the available DSA runs, but a vessel perforation was clearly mentioned by the operator and was therefore included in the analysis. To reduce potential bias towards poor clinical outcome introduced by the moment of classification of the vessel perforation, we repeated the analysis on the association between perforation and clinical outcome with vessel perforations diagnosed by the operator only.

Within the group of patients with vessel perforation, we assessed if perforation location was associated with mRS at three months and NIHSS at 24-hour follow-up using the appropriate regressions mentioned above. In addition, within the vessel perforation group we assessed the association of reperfusion status with mRS at three months and NIHSS at 24-hour follow-up using the same regressions methods. We excluded the four cases in which we could not identify the contrast extravasation on DSA for these subgroup analyses.

Missing data concerning prognostic patient and interventional characteristics were imputed with multivariable imputation by chained equations (MICE) package.^16^ The level of significance was defined as p < 0.05. The analyses were performed in R (version 4.2.3; R Foundation for Statistical Computing, Vienna, Austria).^17^

## Results

We included 2794 patients, of which 74 (2.6%) had a vessel perforation. Supplemental Figure 1 provides information on the amount of vessel perforations scored, and reassessment of DSAs of patients with intracranial hemorrhage on follow-up CT.

Clinical characteristics of patients with and without a vessel perforation are shown in Table 1. Vessel perforation occurred in 14 patients (19%) by a microwire, in 25 patients (34%) by the microcatheter, in 18 patients (24%) by the stent-retriever, and in 6 patients (8%) by contact aspiration. In the remaining 11 patients (15%) the cause of vessel perforation was unknown. We observed 39 (57%) distally located vessel perforations and 30 (43%) proximal vessel perforations. In one patients the location of vessel perforation was unknown. We observed 62 (88%) subarachnoid type, 4 (6%) parenchymal type, and 4 (6%) arterio-venous fistula type contrast extravasations. In four patients the vessel perforation type could not be assessed. After a vessel perforation the procedure was immediately terminated in 25 patients. Subsequent DSA runs without any intra-arterial treatment were performed in 29 patients. In 11 patients EVT continued in another vessel. In 5 patients EVT continued in the same vessel but more proximally. In 4 patients the interventionalist resolved the contrast extravasation by gluing or coiling the particular vessel.

**Table 1.**
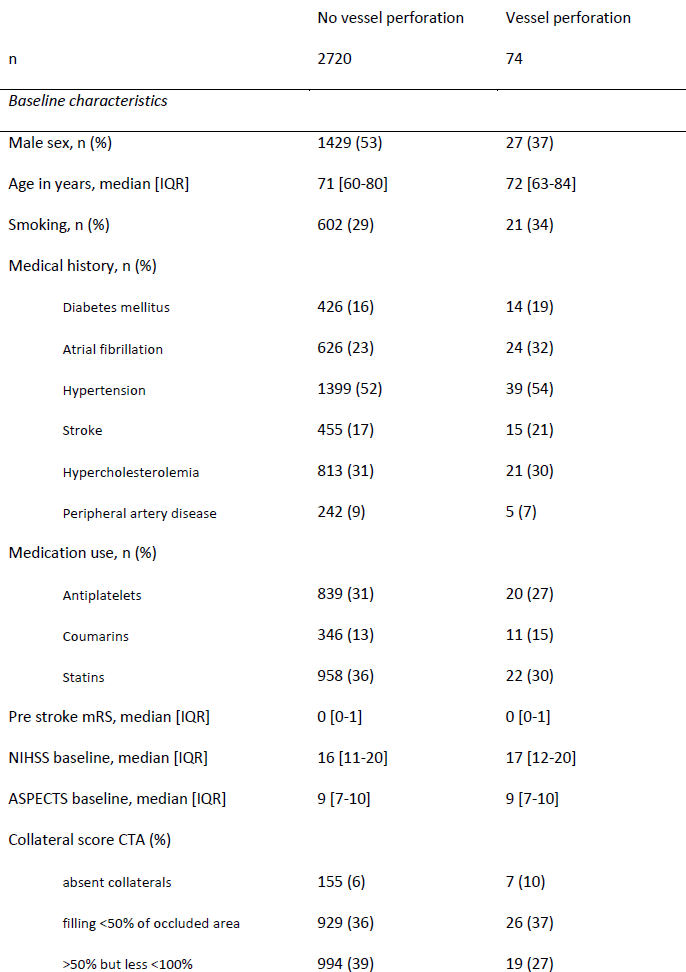
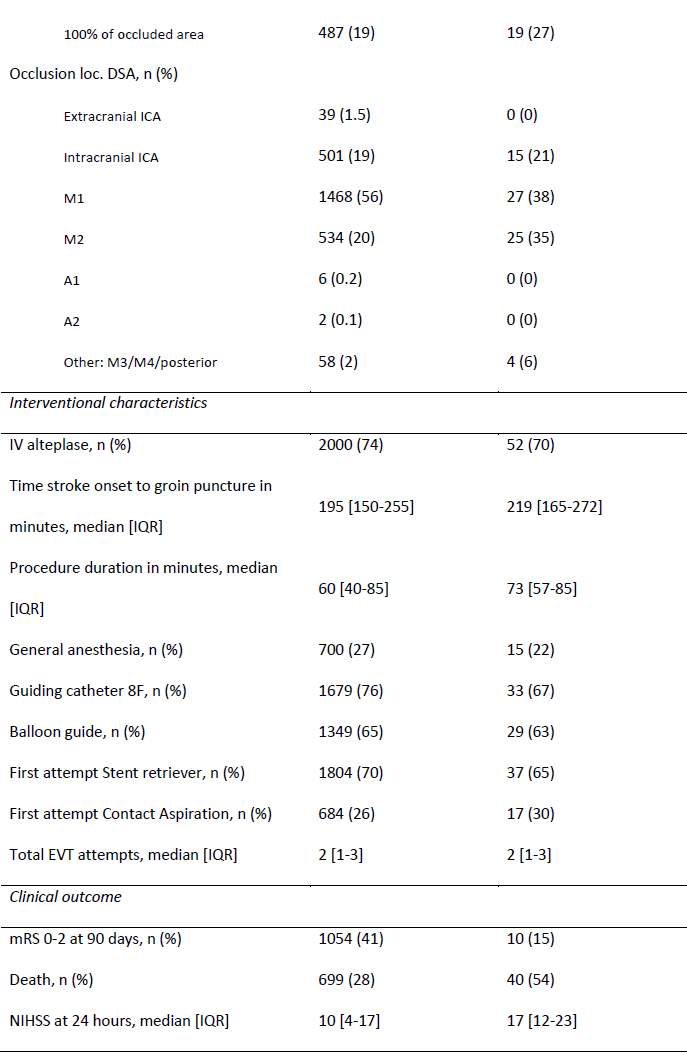
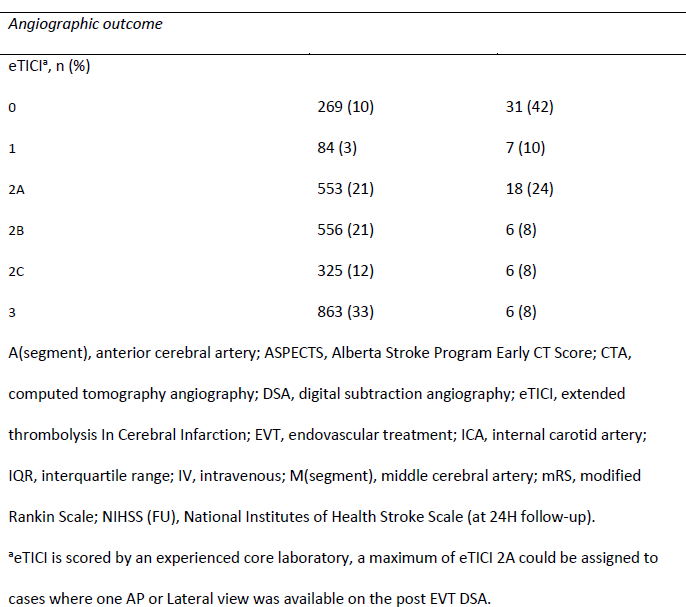
Baseline, interventional and outcome parameters in patients with and without a vessel

### Determinants of vessel perforation

Results of the univariable regression analysis of determinants of vessel perforations can be found in supplementary table 1. The multivariable regression analysis revealed that female sex and distal occlusion locations were associated with a higher odds of a vessel perforation (resp. aOR 1.9 [95%CI 1.2-3.1], aOR 2.2 [95%CI 1.4-3.7]). No statistically significant associations were observed with respect to administration of intravenous thrombolytics (IVT), the time from stroke onset to the start of the procedure, use of general anesthesia or the total amount of EVT attempts. The majority of patients in both groups were treated with a stent retriever first, and no significant difference was found concerning the proportion of using stent retriever or aspiration technique between both groups.

### Association between vessel perforation and outcome

Vessel perforation was associated with worse functional outcome, cOR 0.30(95%CI 0.19-0.48), also after adjustment for patient parameters and TICI score, acOR 0.41 (95%CI 0.25-0.68). Vessel perforation was negatively associated with good functional outcome (aOR 0.40, 95%CI 0.19-0.86) and mortality (aOR 3.2, 95%CI 1.8-5.6). Vessel perforation was associated with higher in-hospital NIHSS at 24-hour follow-up, aβ 0.34 (95%CI 0.16-0.51).

The association between vessel perforation and mRS at 90 days did not change after exclusion of patients in whom vessel perforations were recognized at follow-up CT only (Supplemental table 1).

### Angiographic features of vessel perforation and clinical outcome

Distal perforations were only associated with a shift towards better functional outcome in the unadjusted model, but not in the adjusted model (Table 2). No difference was observed concerning risk of poor outcome at 90 days, mortality and NIHSS at 24H follow-up in patients with distal compared to proximal perforation location.

**Table 2.** Association of location of vessel perforation and successful reperfusion with functional outcome, mortality and NIHSS at 24H.

|  | mRS90d ordinal |  | mRS90d 0-2 |  | Death |  | NIHSS 24H<sup>†</sup> |  |
| --- | --- | --- | --- | --- | --- | --- | --- | --- |
|  | cOR<br>(95%CI) | acOR*<br>(95%CI) | OR<br>(95%CI) | aOR*<br>(95%CI) | OR<br>(95%CI) | aOR*<br>(95%CI) | Beta<br>(95%CI) | Beta*<br>(95%CI) |
| Distal vs Proximal | 3.3 (1.2-9.5) | 2.7 (0.7-10.8) | 3.9 (0.8-18.9) | - | 0.4 (0.1-1.0) | - | -0.3 (-0.6-0.02) | - |
| eTICI ≥2B | 2.2 (0.7-6.2) | - | 2.8 (0.7-11.6) | - | 0.5 (0.2-1.7) | - | -0.5 (-0.8- -0.1) | -0.3 (-0.6-0.01) |

Adjusted regression analyses are not performed when unadjusted regression was not significant.
acOR, adjusted common odds ratio; cOR, common odds ratio; mRS90d, modified Rankin Scale at 90d; NIHSS FU24H, National Institutes of Health Stroke Scale at 24h follow-up
\* model adjusted for: age, pre-stroke mRS dichotomized (0-2), history of stroke, history of Diabetes Mellitus, glucose blood level (mmol/L), systolic blood pressure (mmHg), NIHSS at baseline, collateral grading score dichotomized (≥50%), ASPECTS at baseline, IV- thrombolytics, time stroke onset to groin puncture, occlusion segment proximal/distal on DSA, reperfusion dichotomized (eTICI ≥2B)
† NIHSS at 24H was log transformed to meet the assumptions of normally distributed residuals for linear regression.

Reperfusion was successful in 18/74 (24%) of patients with vessel perforation. No association was found between successful reperfusion and functional outcome or mortality (Table 2). Successful reperfusion was associated with lower NIHSS at 24H follow up in the unadjusted model only (Table 2).

## Discussion

This study investigated vessel perforations during EVT in ischemic stroke patients. We report an incidence rate of 2.6%, in which women and distal occlusion locations were associated with a vessel perforation. Vessel perforation has severe clinical consequences, as we found a two-fold higher risk of poor functional outcome and mortality rate compared to non-perforated cases. Visually, the majority of contrast blushes were of a subarachnoid type. There were slightly more distal vessel perforations than proximal, but clinical outcome was similar in both groups. Successful reperfusion was achieved in a minority of patients with vessel perforation, and did not show an association with functional outcome.

The incidence rate we found is comparable with previous registry studies in which 0.7-2.6% of patients had a vessel perforation.^2,4,10,18-20^ Some studies define vessel perforation as subarachnoid hemorrhage on follow-up CT, with incidence ranging from to 2.9% to 20%.^3,19,21^

Distal occlusion locations and female sex increase the risk of vessel perforation. Smaller vessels have smaller lumens and thinner vessel walls, which could explain the increased risk of vascular damage during EVT.^22^ The association with sex could be explained by the smaller vessel size in women compared to men. Studies measuring intracranial vessel wall thickness and lumen diameter in the circle of Willis on MRA showed thinner vessel walls and lower vessel diameters in females than in men.^23–25^ No literature was found on distal vessel dimensions in relation to sex, however, if proximal vessels are smaller in females, this will probably also be true for distal vessels.

The occurrence of contrast extravasation was most often due to manipulation of microcatheter, followed by stent-retriever and microwire manipulation. Keulers et al. showed reduced vessel perforations when passing the clot using a microcatheter with the microwire retracted 10 mm from the tip, instead of a microwire alone.^19^ In our study we could not study this specific method of clot passing. However, we did find that microcatheter manipulation towards the clot also resulted in a perforation. In a small case study, Matsumoto et al. described an anchor wire technique using a basket-shaped microwire which adheres to the vessel to stabilize the exchange of EVT devices near the target vessel to reduce manipulation induced vessel perforations.^26^

The clinical outcome of vessel perforations is poor. Potentially, the discontinuation of EVT before reperfusion is a major factor leading to worse clinical outcome. In a case report of two patients, interventionists were able to glue a distal M2 vessel perforation and continue with the thrombectomy of a proximal M1 occlusion achieving successful reperfusion. Functional outcome was stated to be good (mRS at 90d ≤ 2) in both cases.^27^ In a recent case series comparing successful and unsuccessful reperfusion in vessel perforation patients, the authors show favorable clinical outcome in patients with successful reperfusion.^10^ This raises the question if the discontinuation of EVT could be the main contributor to worse clinical outcome, instead of the vessel perforation itself. We did not find an association between successful reperfusion and clinical outcome in vessel perforation patients. This finding differs from the study of Ducroux et al and could be explained by the large proportion of ICA occlusions in our study (31% vs 21%) in addition to the larger proportion of unsuccessful reperfusion in these occlusions (37.5% vs 16.7%).^10^ We did find a favorable association of successful reperfusion with NIHSS at 24H follow-up, but failed to show significant association after adjustment for prognostic patient parameters.

The number of EVTs performed for distal occlusions is increasing, from approximately 8% in the EVT trials in 2015 to 15-54% in the latest RCTs on EVT.^5-9,28-34^ Ongoing trials, such as the DISTAL and ESCAPE-MeVO trials, specifically try to estimate the benefit of EVT in these patients (resp. NCT05029414 and NCT05151172). M2 occlusions are considered good EVT targets according to stroke physicians.^35^ In our study, distal occlusions were at higher risk of a vessel perforation, but the clinical outcome of distal perforations was similar to proximal perforations. Potential treatment benefit could be lower in distal occlusion locations due to increased rate vessel perforations. However, a vessel perforation remains a rare complication, and therefore the treatment benefit in distal occlusion patients potentially outweigh the increased risk of a vessel perforation.

### Limitations

A limitation of our study followed the design of the MR CLEAN Registry where follow-up imaging was only performed when a patient clinically deteriorated. Therefore, potential vessel perforations in patients without a decline in neurological performance and missed by operator and core laboratory on DSA could not be assessed. By not recognizing the minimally symptomatic or asymptomatic vessel perforations, the odds for worse outcome could be exaggerated. However, due to the limited amount of cases found in patients with hemorrhage on CT, we believe the amount of missed cases to be small.

The limited amount of patients prevents us from definitively state that there is no difference in clinical outcome between proximal and distal vessel perforations or reperfusion status, although this is the largest cohort of vessel perforations in current literature.

## Conclusion

The incidence of vessel perforation during EVT is low, but it has severe clinical consequences. Females and patients treated at distal occlusion locations are at higher risk. The location of vessel perforation EVT and successful reperfusion did not significantly affect functional outcome in our cohort.

## Data Availability

Data is available through the data-request procedure of MR CLEAN trial office. Contact: Naziha el Ghannouti, research coordinator Erasmus MC University Medical Center Ee 2234 PO BOX 2040 3000 CA Rotterdam The Netherlands Tel: +31 10 704 3818 Fax: +31 10 704 4721

## Nonstandard Abbreviations and Acronyms

A1-A2-A3: – segment of anterior cerebral artery
DSA: – Digital Subtraction Angiography
EVT: – Endovascular thrombectomy
eTICI: – extended Treatment In Cerebral Ischemia score
ICA: – internal carotid artery
IVT: – intravenous thrombolysis
M1-M2-M3: – M1-M2-M3 segment of middle cerebral artery
mRS: – modified Rankin Scale
NIHSS: – National Institutes of Health Stroke Scale

## Acknowledgements

We thank the MR CLEAN Registry investigators for their contribution. A list of all investigators is given in the supplement material.

## Personal disclosures

P. Matthijs van der Sluijs – no conflict of interest.

R. Su – no conflict of interest.

S.A.P. Cornelissen – no conflict of interest.

A.C.G.M. van Es – no conflict of interest.

G. Lycklama a Nijeholt – no conflict of interest

B. Roozenbeek – no conflict of interest.

P.J. van Doormaal – no conflict of interest.

J. Hofmeijer – no conflict of interest.

A. van der Lugt – no conflict of interest.

T. van Walsum – no conflict of interest

---

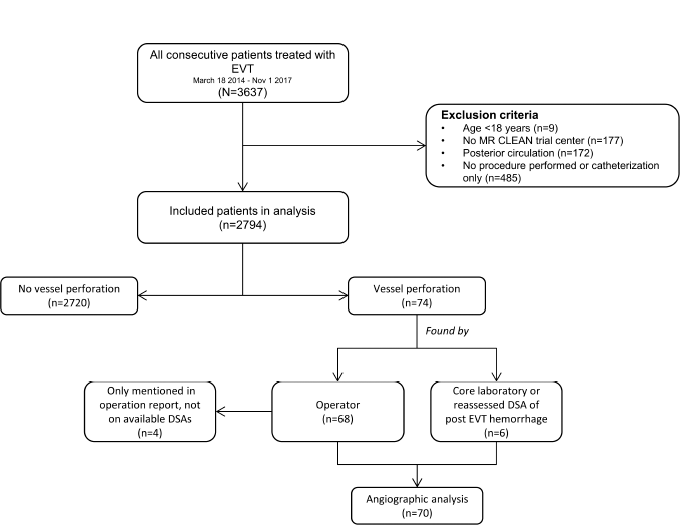

